# Virtual Responsive Neurostimulation Implantation: From Intracranial Connectivity to Optimized Lead Placement

**DOI:** 10.64898/2026.06.17.26355892

**Authors:** Odile Feys, Katherine G. Walsh, Kerry C. Nix, Mariam Josyula, Nishant Sinha, Sarah B. Lavelle, Joost Wagenaar, Andrew Michalak, Martha J. Morrell, Jay Jeschke, Ankit N. Khambhati, Erin C. Conrad, Jonathan K. Kleen, Brian Litt, Vikram R. Rao, Daniel Friedman, Kathryn A. Davis

## Abstract

Responsive neurostimulation (RNS) is an implanted device that delivers direct brain stimulation for drug-resistant focal epilepsy. Individual responses are highly variable, and no validated framework exists to predict outcome or guide lead placement before implantation. We hypothesized that this variability is partly explained by lead placement in relation to patterns of functional connectivity in brain networks.

Fourty-nine patients with drug-resistant focal epilepsy who underwent pre-implantation intracranial EEG (iEEG) and RNS implantation across three independent epilepsy centers were retrospectively studied. We developed a composite functional connectivity score, based on simple Spearman correlation, combining the standard deviation and kurtosis of interictal iEEG connectivity distributions to predict the response outcome in a training cohort (HUP, n=18) and validated in two independent cohorts (NYU, n=17; UCSF, n=14). We accounted for a spatial mismatch between iEEG and RNS electrodes with a distance-based correction. The score was extended to generate patient-specific 3D maps of predicted RNS efficacy across 200 simulated, or “virtual RNS”, lead configurations.

Accuracy of the score in predicting clinical outcome was 72% at the group level, 61% at the individual patient level, and, after distance-based optimization, 100% in patients with RNS electrodes placed close to location of iEEG electrodes. Applied to the validation cohort, the same score reached 68% accuracy (71% balanced accuracy, 55% sensitivity, 88% specificity). The spatial combination of the scores at different SEEG contacts localization gives a spatial score for each patient. Responders showed significantly higher spatial scores than non-responders, supporting that actual RNS lead placement in responders was located in map-identified favorable regions.

Interictal iEEG functional connectivity predicts individual RNS response across independent epilepsy centers, and patient-specific 3D maps derived from this biomarker could prospectively guide lead implantation toward favorable network regions, opening a promising avenue toward network-informed RNS surgical planning.

## Introduction

Despite advances in antiseizure medications, approximately one-third of patients with epilepsy remain drug-resistant to pharmacological treatment, and a substantial proportion are not candidates for resection epilepsy surgery, e.g., because the epileptogenic zone (EZ) cannot be reliably removed ^1^. In these patients, neurostimulation has emerged as a therapeutic alternative, and responsive neurostimulation (RNS) has demonstrated sustained seizure reduction in multicenter trials over long-term follow-up ^2–5^.

However, RNS outcomes are highly variable across individuals. While a majority of patients achieve meaningful seizure reduction, a third derive limited benefit ^6^, and the factors explaining this heterogeneity remain poorly understood ^7^. Identifying reliable biomarkers of RNS response before implantation would improve patient selection and surgical planning ^5^. Existing approaches have predominantly shown group-level differences in responders vs non-responders, but no accurate prediction of the individual patient’s response. This lack of predictive biomarkers at a patient-level limits individual patient decision-making.

Beyond patient selection, the location of RNS lead placement is a critical and largely unresolved determinant of therapeutic efficacy ^8^. Lead placement is currently guided by the presumed EZ localization based on non-invasive assessment and intracranial EEG (iEEG) recordings ^9^, but no validated framework exists to identify optimal implantation sites from a network perspective ^10^. Importantly, this EZ-centered approach may not be the optimal approach or even excludes patients in whom the EZ cannot be precisely delineated. For these patients, RNS represents a palliative option whose efficacy may depend critically on assumptions for lead placement, yet current practice offers no established data-driven guidance for this decision ^11^.

Functional connectivity, i.e., temporal correlations between spatially remote neurophysiological signals ^12^, measured from iEEG recordings offers a promising substrate for this purpose since global synchronizability moderately discriminated responders and non-responders ^13^, as it captures the dynamic organization of epileptic networks. It is agnostic to the EZ localization, and is available in all candidates undergoing invasive monitoring.

In this multicenter study, we aim to develop and validate a functional connectivity biomarker predictive of individual RNS response. We further demonstrate that this biomarker can be translated into patient-specific 3D maps of expected RNS efficacy to prospectively guide lead implantation.

## Methods

### Patients

Patients with drug-resistant focal epilepsy from three university-based comprehensive epilepsy centers (Hospital of University of Pennsylvania (HUP), New York University (NYU) Langone and University of California San Francisco (UCSF)) were retrospectively included in this study. All the patients were older than 18 years and underwent iEEG prior to RNS device implantation with a minimum two-year follow-up. Patients were classified according to their clinical (self-reported) seizure frequency at two years after implantation, with responders defined as patients with at least 50% clinical seizure frequency reduction relative to the baseline. The study was approved by the ethics committee (common IRB 829175) and patients provided written informed consent to be included in the study.

### Signal acquisition and preprocessing

#### Intracranial electroencephalography

Implantation scheme (Figure 1) was based on anatomo-electrico-clinical hypothesis of the epileptogenic zone localization following non-invasive assessment. At HUP, 83% of depth and 17% of subdural electrodes were implanted (Ad Tech Medical Instruments), and signal was recorded at 512 or 1024Hz (Natus Quantum amplifier, high-pass filter: 0.08Hz, antialiasing low-pass filter: 219-439Hz). At NYU, 41% of depth and 59% of subdural electrodes were implanted (Ad Tech Medical Instruments and Dixi), and signal was recorded at 2048Hz (Natus Quantum amplifier). At UCSF, 30% of depth and 70% of subdural electrodes were implanted (Ad Tech Medical Instruments), and signal was recorded at 512 or 1024Hz (Natus Quantum amplifier). To harmonize iEEG data across centers, downsampling to 512Hz was applied with no additional filters.

**Figure 1.**
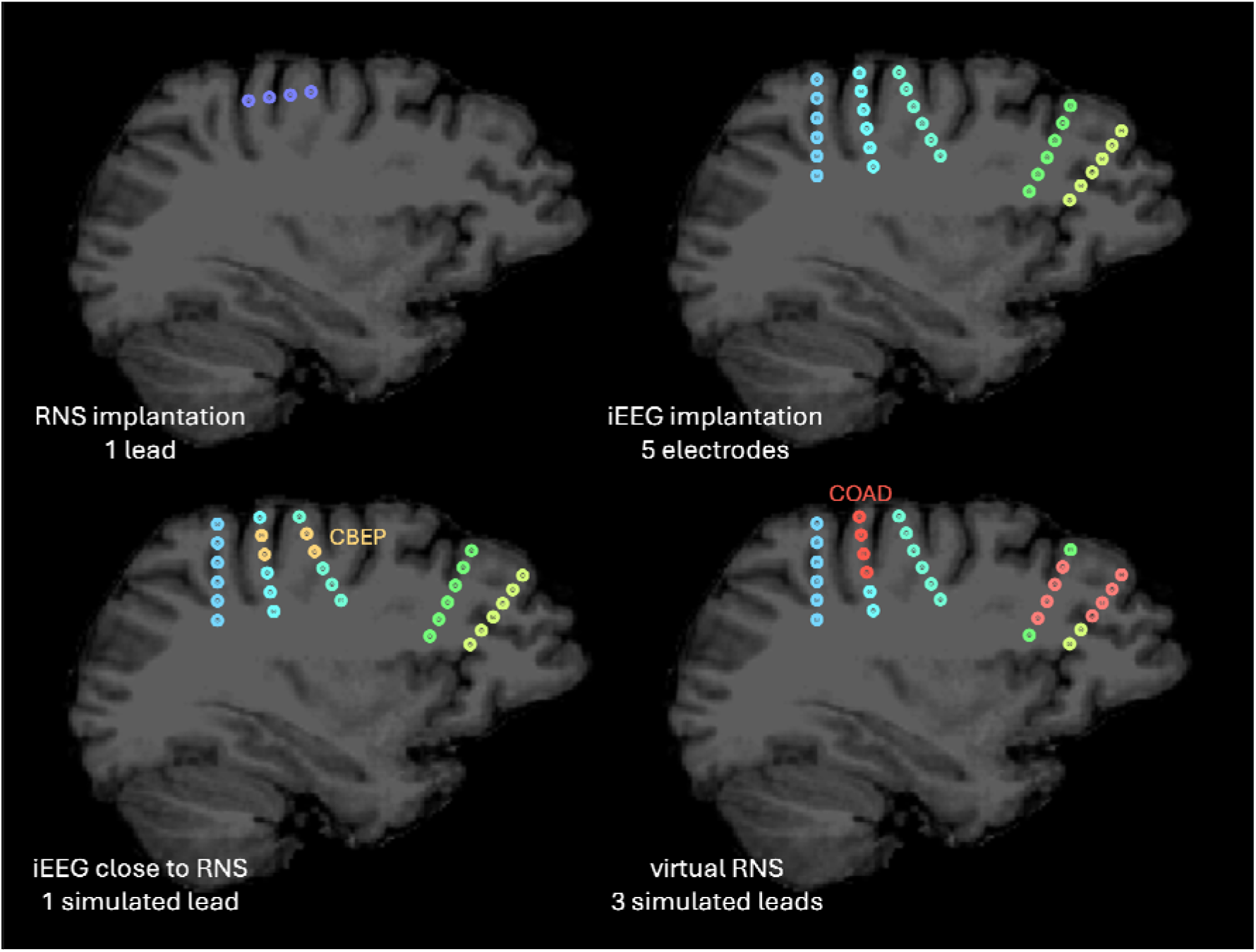
Schematic representation of virtual RNS implantation. **Top left.** Actual RNS implantation with one implanted lead (dark blue). **Top right.** Actual iEEG implantation with five implanted electrodes (cyan, light blue, turquoise, light green, yellow). **Bottom left.** Selection of the iEEG contacts as close as possible to the RNS implantation (i.e., the combination with the best electrode proximity, CBEP, orange). **Bottom right.** Virtual RNS implantation with simulation of three RNS leads (dark and light red) based on iEEG implantation (the closest to the actual RNS implantation is named the combination with optimal assignment distance, COAD, dark red). For illustrative purpose, the schematic representation shows an example of one of the two implanted RNS leads (top left) and three virtual RNS implantations out of the 200 generated random combinations (bottom right).

#### Responsive neurostimulation

Two to five leads were implanted per patient, with two of these leads connected to the RNS device (RNS-320, NeuroPace, Inc, Mountain View, CA, USA) across the three centers. Implantation scheme was defined based on the consensus of each center’s multidisciplinary epilepsy care team during epilepsy surgical conference. Recording (digitized at 250 Hz per manufacturer settings) and stimulation were performed through two four-contact leads, i.e., four pairs of contacts in bipolar montage, according to respective institutional practices.

#### Localization of intracranial contacts

Precise localization of each iEEG and RNS contact was determined using in-house software of each center applied to linearly registered preimplantation and postimplantation T1-weighted brain MRI and postimplantation brain CT, previously described in ^14^. Based on the known localization in the same three-dimensional space, Euclidean distance between iEEG and RNS contacts was computed.

#### Distribution of cohorts

Hereafter, the HUP patients served as the training cohort, whereas the NYU and UCSF patients comprised the validation cohort.

### Preliminary checks on the training cohort

Prior to developing an iEEG connectivity-based biomarker of RNS response, we first assessed whether connectivity warranted further investigation as a candidate biomarker ^13^. We hypothesize that if local-scale, i.e., restricted to the area surrounding RNS leads, connectivity within the implanted region does not differ between groups, functional connectivity is unlikely to be the optimal biomarker for predicting RNS response. In contrast, if RNS-measured local-scale connectivity differs between responders and non-responders, it will confirm our interest in local intracranial functional connectivity to develop a biomarker. Because exact localization of iEEG electrodes and RNS leads can differ within a patient and result in different signals/connectivity patterns, the RNS signal was used as the ground-truth to build the biomarker, to demonstrate whether or not different local connectivity should be further used as a biomarker to distinguish responders and non-responders and to assess individual response.

For that purpose, we verified whether connectivity analyzed from RNS electrodes differed between responders and non-responders, and that stimulation induces connectivity changes across implantation in our training cohort.

#### RNS signal processing

We used the recorded RNS signal of the first 10 clips with long episodes (early window) and the last 10 clips with long episodes (late window) detected across the two-years’ timeframe. Long episodes are those detected as abnormal/epileptic activity by the RNS device. We selected those long episodes instead of scheduled episodes because the latter are overwritten when several long episodes are detected (in some patients no scheduled episodes were recorded before long episodes). The selection of scheduled episodes would have led to a broader event selection window, with a variable number of intracranial stimulations ^15^ occurring between episodes, potentially leading to variations in local network connectivity depending on stimulation in each patient. Given their limited predictive value (∼34%) for clinical seizures ^16^, long episodes were considered as predominantly interictal activity for the purpose of this study, and most of possibly remaining ictal activity is removed by preprocessing described below. Each clip around the detected long episode represents 90 seconds of bipolar signal which is used for further analyses. Mean and standard deviation (SD) of the signal were computed on the first second of each recorded intracranial clip and used as a reference to identify artifacts. To remove stimulation artifacts and signal saturation (e.g., ictal signals), any timepoint within an clip that exceeded mean ± 5 SD of the signal was excluded from analysis ^17^. Connectivity was subsequently computed on the remaining timepoints of each clip. Pairwise functional connectivity was computed between all RNS electrode pairs (as previously described for iEEG functional connectivity analyses ^18^) using Spearman correlation ρ values, a non-parametric metric robust to outliers and amplitude variations, and capable of capturing nonlinear relationships between EEG signals ^19^. Connectivity was computed on the raw voltage signal, providing a frequency-agnostic measure that captures co-variations across the full spectral content without requiring an a priori selection of frequency bands, though this measure is more sensitive to lower-frequency content due to the 1/f structure of the signal.

Correlations with absolute ρ value below 0.05 were considered negligible and interpreted as reflecting absence of functional connectivity between the implied contacts, consistent with established thresholds for trivial effect sizes ^20^.

#### Group-level analyses

Kernel density estimate (KDE) was applied to evaluate the distribution of connectivity values (across all timings, all connections, and all subjects) in responders vs. non-responders in the early window. The distributions were compared using L1 distance (i.e., sum of absolute differences of vector space coordinates), and statistical significance was established using a two-sided maximum statistic ^21^ with 10,000 permutations between labels for each patient with no independence assumption across electrode pairs and timings. Based on these results, early and late windows were then compared either across all patients pooled together (if no significant difference was found between responders and non-responders) or separately within each group (if a significant difference was found).

### Single-center biomarker development

To account for the spatial specificity of RNS recordings, pairwise functional connectivity was computed between the iEEG contacts located as close as possible to the RNS leads.

#### iEEG signal processing

Five-minute interictal iEEG signals were extracted from iEEG recordings (three to five segments per patient, depending on the provided and anonymized iEEG signals, selected at least two hours from seizures to avoid pre- or post-ictal signals). Interictal segments were utilized to be comparable to the RNS long episodes due to the high number of non-ictal detections (at least not related to clinical seizures) inducing intracranial stimulation in patients with implanted RNS ^22^. Four pairs of intracranial contacts were selected based on the Euclidean midpoint of each pair, i.e., those closest to the RNS pairs of contacts, and this combination was referred to as the combination with the best electrode proximity (CBEP; Figure 2 top middle). The iEEG signal was divided in 30-second windows, i.e., the minimal amount of data used in the previous RNS analyses since up to two thirds of data may be excluded by artifact rejection. To increase the number of available windows, the first and last 30-second epochs were retained and the intervening 4-minute segment discarded to avoid inclusion of non-independent consecutive epochs. Spearman correlations were applied between the pairs of electrodes in the CBEP (similar to RNS signals).

**Figure 2.**
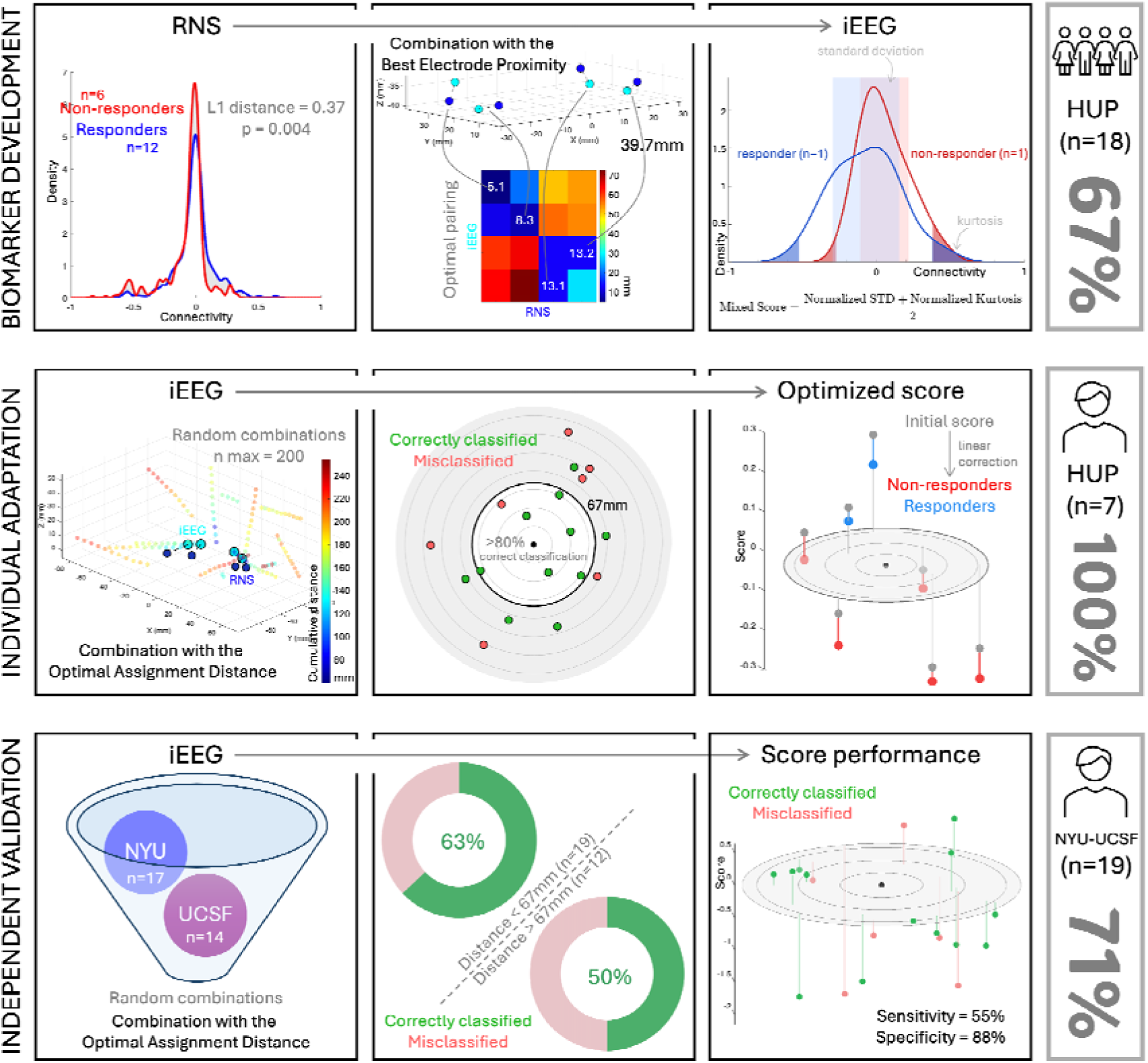
Development and validation of the connectivity biomarker for RNS response prediction. **Biomarker development** (HUP training cohort, n=18). **Left**. Connectivity distributions differ significantly between responders and non-responders in the early RNS window (L1 distance = 0.37, p = 0.004). **Middle**. iEEG contacts are selected based on their proximity to RNS leads (combination with the best electrode proximity, CBEP; shortest cumulative distance = 39.7mm in the illustrated patient). **Right**. The composite score combines the standard deviation and kurtosis of the individual connectivity distribution to classify responders and non-responders; one responder and one non-responder are displayed for the example (group-level balanced accuracy = 72%). **Individual adaptation** (HUP training cohort, n=18 reduced to n=7). **Left**. 200 random iEEG contact combinations are generated to mimic RNS lead geometry, and the combinations and selection of the combination with the optimal assignment distance (COAD). **Middle**. The maximal distance threshold of 67mm is derived from a logistic regression model to retain only patients with a predicted probability of correct classification above 80%. **Right**. A centered linear distance correction is applied to the composite score, achieving 100% balanced accuracy (sensitivity and specificity = 100%) in patients below the distance threshold. **Independent validation** (NYU and UCSF cohorts, n=31 reduced to n=19 with COAD-RNS distance below 67mm): **Left**. The same pipeline is applied without retraining to the two independent validation cohorts combined. **Middle**. Correct classification is significantly more frequent below the distance threshold than above (63% vs. 50%). **Right**. The optimized score achieves 71% balanced accuracy (with 55% sensitivity and 88% specificity) in the validation cohort.

#### Patient-level analyses

To enable single-patient prediction and to capture the subtle variations related to heterogeneity across patients, the KDE distribution was collapsed into a composite score combining the standard deviation and kurtosis of the connectivity value distributions for each patient. The standard deviation captured the spatiotemporal variability of functional connectivity, consistent with its established use in non-invasive connectivity studies ^23^, and the kurtosis captured deviations from normality in the connectivity distributions, reflecting the non-Gaussian structure of resting-state iEEG signals ^24^. Both metrics were assigned equal contribution to the composite score, as it yielded optimal balanced accuracy in the training cohort. The raw features could not be used in the score because of the differences in ranges related to the features, each feature was subsequently z-scored across the group of patients, and both features were summed. For ease of interpretation, the signs of all features were inverted so that higher values corresponded to responders, and a zero threshold was used to classify responders and non-responders (i.e., positive values predicted responders and negative values predicted non-responders). Performance of the score was then assessed in the training cohort.

### Adaptation for individual patient use

After constructing a metric capable of classifying patients as responders or non-responders at the group level, we aimed to adapt it for individual-patient prediction, which is more representative of a potential future clinical use. For that purpose, normalization was required at the patient level.

#### iEEG contacts selection

To simulate RNS electrode configurations from iEEG data (virtual RNS, Figure 1), among all possible combinations of intracranial contacts, 200 combinations of four pairs of contacts were selected based on the following criteria: 4 consecutive contacts on two single probes (i.e., the main difference with the CBEP selection) with an intercontact distance <15mm, yielding four bipolar signals across two electrodes, thereby mimicking the geometric configuration of RNS leads (in contrast with the CBEP selection) and excluding non-spatially adjacent contacts on grids. This number (200) was chosen to balance adequate spatial coverage of the iEEG implantation with computational feasibility in a clinical setting. These 200 virtual couples of RNS leads (random combinations) were balanced across electrodes using round-robin scheduling to ensure adequate representation of underrepresented combinations. Optimal pairing between each iEEG combination and the RNS contacts was defined by the shortest cumulative distance, i.e., the sum of the four distances between each iEEG-RNS pair.

#### Individual patient adaptation

The same processing as previously described (see *iEEG signal processing*) was applied to each combination. The standard deviation and kurtosis were computed on connectivity values for each combination. The combination with optimal assignment distance (COAD; i.e., with the shortest cumulative distance compared to RNS electrodes; Figure 2 middle left) was selected to classify between responders and non-responders. The COAD standard deviation and kurtosis were z-scored with other combinations selected in the same patient. The composite score and classification were applied on the COAD as previously described (see *Patient-level analyses*). Performance of the score was then assessed in the training cohort.

#### Optimization for distance between iEEG and RNS contacts

Both CBEP and COAD were selected based on their proximity to the RNS leads, but a perfect overlap is not realistic. Additionally, the geometric constraint applied to define COAD should result in a larger COAD-RNS distance than the CBEP-RNS distance. As COAD is more representative of simulated RNS implantation than CBEP, COAD will be used for further analyses. Based on this observation, logistic regression was used to model the probability of correct classification (responder vs. non-responder) based on individual analysis according to the COAD-RNS distance. From this logistic function, a maximal distance threshold was computed to maintain the predicted probability of correct classification over 80% (Figure 2 middle middle). All the patients whose COAD exceeded the threshold were discarded from further analyses, leading to the exclusion of patients with a high distance between simulated and actual RNS leads for which the model would not be accurate. Based on the assumption that, within the remaining sample of patients, distance might still impact the performance of the score, a distance correction was expected to improve the score performance. The composite score was adjusted using a centered linear distance correction which decrease the good outcome probability according to the distance, i.e., corrected-score = score - α × (d - δ) / σ where α is a dimensionless weight controlling the strength of the correction, d is the patient-specific COAD-RNS distance, δ is the reference distance, and σ is the scale factor. All combinations of correction were tested with δ ranging from 0 to 65mm in 5mm increments covering all observed COAD-RNS distances in the dataset, σ ranging from 0 to 65mm in 5mm increments, and α ranging from 0 (i.e., no correction) to 0.5 in 0.05 increments. The best correction was determined based on the balanced accuracy, providing equal weight to each class regardless of sample size and class imbalance. In case of equal balanced accuracy, the lowest δ and α and the largest σ were selected to favor spatial similarity between iEEG and RNS implantation and minimize the correction (Figure 2 middle right). Final performance of the corrected score was then assessed in the training cohort.

### Multicentre validation on independent patients

We applied the same individual patient analyses (see *Adaptation for individual patient use*) to the NYU and UCSF cohorts (validation cohorts) using the parameters derived from the HUP training cohort (including optimization for distance) without retraining. Final performance of the score was then assessed in the validation cohort. To verify that optimization for distance was overall relevant, a point-biserial correlation between COAD-RNS distance and classification status (correct vs. incorrect) was computed, and 10,000 shuffled permutations between classification status were applied to assess statistical significance (95^th^ percentile). Percentage of each classification status below and above the distance threshold (defined earlier in HUP) were reported for descriptive purposes.

### Optimal localization of RNS leads

At the current stage, the score is based on the known localization of the actual RNS. This computation is therefore performed after the localization has been determined, i.e., a posteriori approach. However, the score can also be computed for all random combinations of iEEG channels and subsequently used to identify the optimal localization, i.e., a priori approach.

#### Patient-specific spatial score construction

For this a priori approach, the same composite score was computed for each iEEG random combination (including leave-one-out normalization, i.e., z-score of the combination using all the other combinations of the selected patient). Each combination represented a simulated RNS placement with two leads and 4 contacts on each lead. The centroid of each simulated lead was projected in the patient-specific three-dimensional space. A 3D KDE map (Gaussian kernel with SD=8mm) was constructed based on the centroids of each combination. Each combination contributed two Gaussian kernels (one per lead centroid) weighted by the sign of its score (+1 if positive, -1 if negative). The resulting 3D map enabled us to interpolate a score at the localization of the four pairs of contacts of the actual RNS leads. Consequently, a region appears as favorable (positive density, displayed in green) if it is more frequently covered by positively-scored combinations than negatively-scored ones, regardless of the magnitude of individual scores. In patients where most combinations share the same score sign the resulting map may appear uniformly green or red, reflecting a genuine dominance of one class of configurations rather than a visualization artifact. The four resulting scores were summed to obtain a single score for each patient. To validate this method, patient scores were compared to assess whether responders had higher scores than non-responders. Since the scores are highly dependent on the implantation strategies across centers, an intra-center z-score normalization was applied. The minimal distance to consider proper coverage of a pair of contacts was set at one-fourth (i.e., ratio defined based on the cumulative distance over the 4 pairs of electrodes and reported to 1 pair of electrodes) of the previous distance threshold and patients with the four pairs of contacts exceeding this minimal distance with at least one centroid were discarded from the comparison. The difference in median score between responders and non-responders was assessed statistically using 10,000 shuffled permutations (significance at the 95^th^ percentile). We considered this score, hereafter referred to as the spatial score, clinically relevant if those of responders significantly exceeded those non-responders.

#### Comparison to the seizure-onset zone localization

The coordinates of the seizure onset zone (SOZ) contacts, as defined by iEEG evaluation, were interpolated onto the KDE map using trilinear interpolation. For each SOZ contact, a normalized density value was extracted. A contact was classified as lying within a favorable region if its density exceeded 0, and within an unfavorable region otherwise.

At the patient level, the SOZ was considered to be predominantly in a favorable region if at least 50% of its contacts had a positive density value. The proportion of patients meeting this criterion was compared against a null hypothesis proportion of 50% (representing random spatial overlap between the SOZ and favorable KDE regions) using a one-sided binomial test.

## Results

Forty-nine patients were included across the three centers: 18 from HUP (training cohort, mean age: 41y, range: 22-61y), 17 patients from NYU and 14 from UCSF (validation cohorts, mean age: 34y, range: 18-62y and mean age: 35y, range: 22-52y, respectively). Across the three cohorts, 31 patients were classified as responders (HUP, n=12; NYU, n=9; UCSF, n=10) and 19 as non-responders at two-year follow-up.

Preliminary RNS analyses confirmed that functional connectivity was a meaningful target for biomarker development (Figure 2 top left). In the early window (first 10 long episodes following device implantation), connectivity distributions already differed significantly between responders and non-responders (L1 distance = 0.37, p=0.004), supporting the biological relevance of this signal. Longitudinal analyses further revealed that this difference was driven by non-responders, whose connectivity distributions changed substantially over time (L1 distance = 0.56, p=0.003), while those of responders remained stable (L1 distance = 0.27, p=0.15), suggesting that successful stimulation may stabilize network dynamics not depending on the implanted brain region.

Building on these observations, we developed a composite connectivity score combining the standard deviation and kurtosis of the connectivity distribution. Applied to iEEG contacts selected for their proximity to the RNS leads (CBEP), the score achieved 72% accuracy (67% balanced accuracy) at the group level. Adapting this approach to individual standalone patient use with virtual RNS implantation (i.e., results for COAD normalized with other simulated leads), a prerequisite for clinical translation, yielded 61% accuracy (67% balanced accuracy), reflecting the added challenge of patient-level normalization in the absence of a reference group.

To address the spatial mismatch inherent to comparing iEEG and RNS recordings, we introduced a distance-based correction. Within the subset of patients whose iEEG contacts fell within a maximal distance threshold of 67mm cumulative across the 4 pairs of contacts, based on the logistic regression, from the RNS leads (n=7 in the training cohort, 2 responders and 5 non-responders), accuracy reached 100% after centered linear distance correction, consistent with the threshold selection criterion designed to retain only patients with a predicted probability of correct classification above 80% (sensitivity and specificity = 100% with α = 0.05, δ = 10mm, σ = 10mm). This strong spatial dependency was confirmed in the validation cohort, with 63% of correct classification below the threshold (n=19, 11 responders and 8 non-responders) versus 50% above (n=12, 8 responders and 4 non-responders).

Applied to the independent validation cohort (NYU and UCSF), the score (without any retraining) achieved 68% accuracy (71% balanced accuracy), with 55% sensitivity and 88% specificity, confirming its generalizability across centers. With no distance correction, the score would have achieved 63% accuracy (63% balanced accuracy) on this cohort, with a chance level of balanced accuracy at 50%. Given the limited sample size within each center (n=8 and n=11), center-specific analyses would be statistically underpowered and prone to spurious differences.

Finally, to translate these findings into a practical surgical planning tool, we extended the composite score to map optimal RNS lead placement at the individual patient level. For each patient, the score was computed across 200 simulated lead configurations derived from iEEG contacts, and the resulting values were projected onto a patient-specific 3D map using KDE. This map provides a continuous spatial representation of predicted RNS efficacy (Figure 3), allowing identification of brain regions where implantation is most likely to yield a therapeutic response (Figure 4).

**Figure 3.**
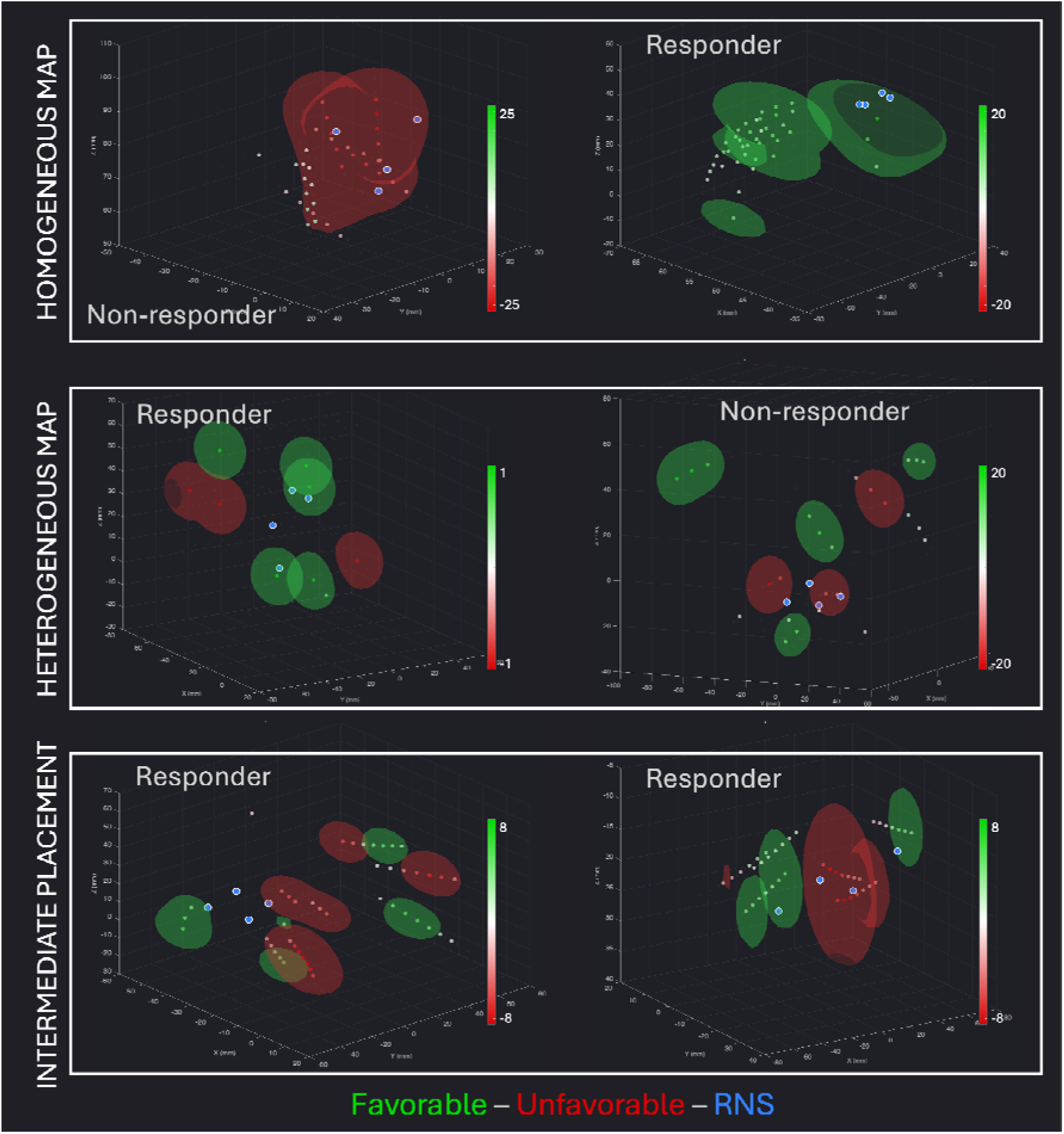
Patient-specific 3D maps of spatial score of predicted RNS efficacy. For each patient, the connectivity score was computed across 200 simulated lead configurations and projected onto a patient-specific 3D map using kernel density estimation. Green and red regions indicate favorable and unfavorable predicted implantation sites, respectively. Each point represents the centroid of a simulated lead (average position of 4 consecutive contacts), not an individual iEEG contact. The spatial distribution and density of points therefore reflect the iEEG implantation scheme, electrode type, and the geometric constraints applied to select valid combinations (4 consecutive contacts on a single probe, intercontact distance <15mm), rather than the full iEEG coverage. Blue dots indicate actual RNS lead locations (one point per pair of contact). Cases are categorized for illustrative purposes based on visual inspection of the map and RNS lead localization. **Homogeneous map**. The map is uniformly favorable or unfavorable regardless of lead placement. **Heterogeneous map**. The map shows distinct favorable and unfavorable regions, illustrating how lead placement determines predicted outcome. Notably, the non-responder (right panel) shows clearly identifiable favorable regions that were not targeted by the actual implantation. **Intermediate placement**. RNS leads are located at the boundary between favorable and unfavorable regions.

**Figure 4.**
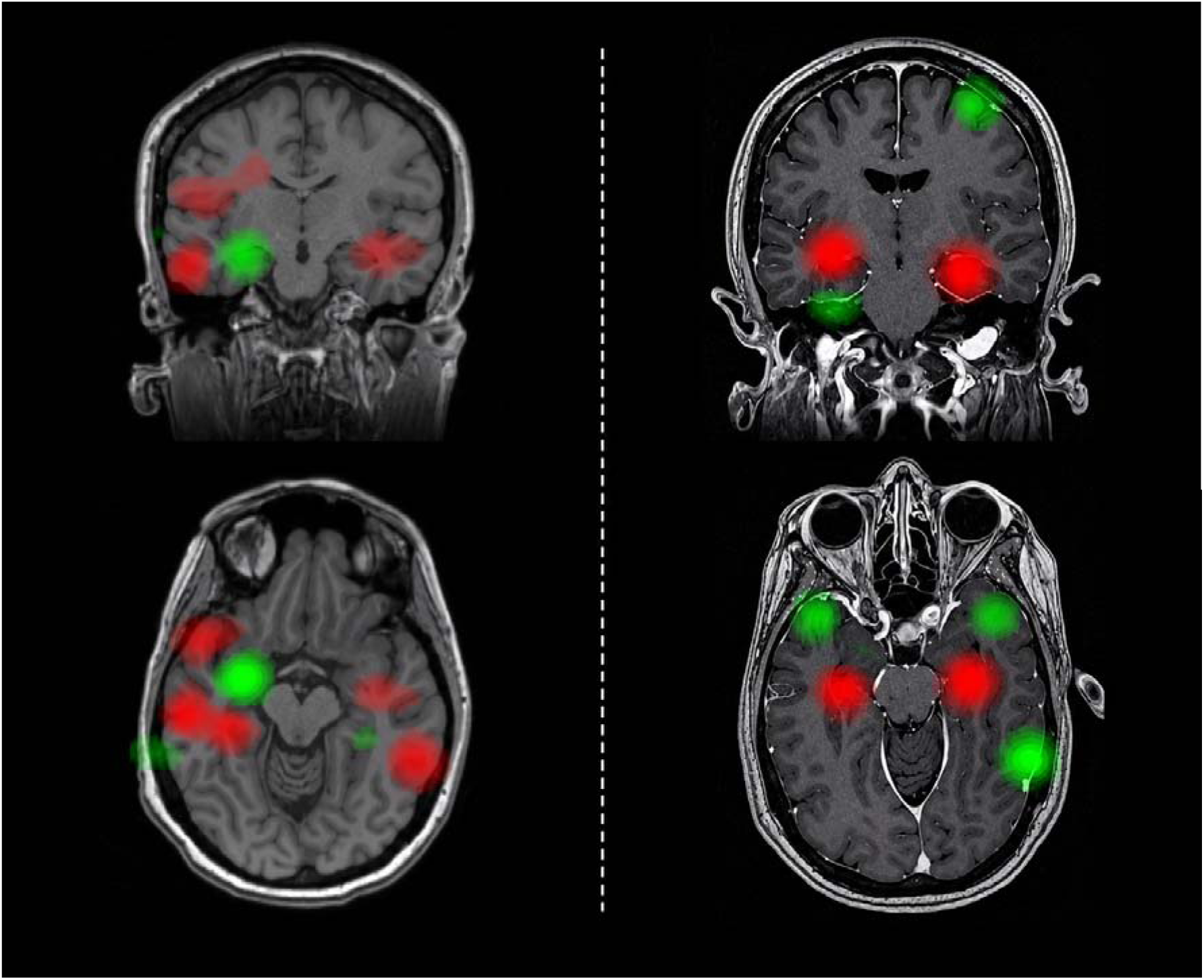
Patient-specific spatial score maps projected onto individual MRI. Two examples of patient-specific spatial score maps projected onto coronal and axial slices of individual patient MRI (radiological convention) using the SPM software^35^. **Left.** A patient with right hippocampal implantation identified as favorable, while neocortical regions appear unfavorable. **Right.** A patient with bilateral hippocampal implantation predicted to be unfavorable, whereas the temporal poles, the right temporal base, and the posterior lateral temporal cortex are identified as potentially favorable targets.

To validate this approach, the spatial score interpolated at the true RNS lead locations was compared between groups. Responders showed significantly higher spatial scores than non-responders (difference of medians = 0.50, p=0.03), confirming that actual lead placement in responders was indeed located in more favorable regions by the map (n=47, after exclusion of 2 patients whose RNS leads were located beyond the per-contact distance threshold, i.e., due to thalamic RNS implantation and no thalamic iEEG implantation). This result suggests that the 3D connectivity map could prospectively guide RNS lead implantation to maximize the likelihood of therapeutic response.

SOZ was not delineated in 5 of the 49 patients (3 responders, 2 non-responders) and these patients were therefore excluded from this analysis. Among the remaining 44 patients, the SOZ was predominantly located within favorable KDE regions in 35 patients (80%, including 22 responders and 13 non-responders), a proportion significantly exceeding chance level (binomial test, p=0.0001). Notably, this finding was independent of treatment outcome, as favorable SOZ localization was observed in 22/28 responders (78%) and 13/16 non-responders (81%).

## Discussion

In this multicenter study, we developed and validated a functional connectivity biomarker derived from interictal iEEG recordings that predicts individual response to RNS. We further demonstrated that its performance is critically dependent on the spatial proximity between iEEG and RNS electrodes, a finding we leveraged to construct patient-specific 3D maps of predicted RNS efficacy that could prospectively guide lead implantation.

### Need for a biomarker

Among multiple tested biomarkers of RNS response, network metrics have emerged as promising candidates supported by the epileptogenic network concept underlying focal epilepsy ^25^. Specifically, previous studies demonstrated that connectivity measures may be relevant: First, network synchronizability computed from ictal iEEG recordings, in a cohort with partial overlap with the present study, predicted RNS response ^13^. Second, effective connectivity computed from cortico-cortical evoked potentials associated with network nodes determined therapeutic outcome ^8^. Our score and its derived 3D map uniquely address these two objectives within a coherent and clinically actionable framework. Additionally, the computation underlying our composite score is strongly supported by non-invasive studies using mean and standard deviation of connectivity distributions based on magnetoencephalography recordings to predict outcome at the group level ^23^. The relevance of lead localization relative to network organization is further supported by studies showing that connectivity between stimulation sites and disease-specific networks predicts seizure reduction ^26^, and that state-dependent effects of RNS differ according to seizure localization ^7,27^. Importantly, patient-specific structural connectivity informed RNS outcomes in mesial temporal lobe epilepsy whereas normative connectivity did not ^28^, underscoring the need for individualized rather than population-based network approaches.

### Biological rationale

The different RNS connectivity distributions between responders and non-responders in the early window suggests that conceptually different networks may be stimulated in both groups. We hypothesized that the stimulated network in responders is intrinsically stable and reinforced, rather than reshaped, by intracranial stimulation, while unstable networks, subject to substantial reorganization driven by further ictal activity, are present in non-responders. This interpretation is supported by evidence that network organization within and outside the EZ differs ^29^ and evolves over the course of the disease ^17,30^, suggesting that epileptic networks are not static but can be steered in different directions, either toward pathological reorganization by recurrent seizures or toward normalization by targeted stimulation. Indeed, long-term analysis of RNS recordings demonstrated that therapeutic benefit is associated with progressive reorganization of interictal functional connectivity over months to years of stimulation ^31^ instead of acute interruption of individual seizures ^32^. Furthermore, stimulation delivered during low seizure risk was previously related to seizure reduction ^33^, supporting the notion that network stability at baseline may favor effective stimulation.

#### Future clinical use

The 3D connectivity map derived from interictal iEEG recordings offers a promising framework for integrating network-based information into RNS surgical planning. In current practice, lead placement is determined by the multidisciplinary epilepsy team based on the presumed localization of the EZ and anatomical constraints ^9^. While SOZ-guided implantation incidentally targets favorable network regions in approximately 80% of patients, the similar overlap observed in responders and non-responders confirms that SOZ localization alone is insufficient to predict individual response, does not capture the full extent of potentially favorable targets identified by the connectivity map, and excludes by definition patients without recorded seizures. The map could serve as a visualization tool, allowing the surgical team to assess whether a planned implantation targets favorable network regions and to adjust lead placement accordingly, and/or as a guide for the final implantation trajectories, e.g., in patients where multiple implantation strategies are equally plausible from an anatomical standpoint, the map could provide an objective, data-driven criterion. Figure 3 illustrates one example (heterogeneous map, right) of a non-responder in our cohort who shows clearly identifiable favorable implantation of brain areas which were not targeted by the actual RNS leads, suggesting that the map-guided placement might have improved therapeutic outcome in this patient and one example (homogeneous map, left) of a non-responder in our cohort who would not benefit at all from RNS and should pursue other treatment strategies. Importantly, this approach is applicable to all RNS candidates who underwent iEEG recording for diagnostic purposes. A prospective validation will ultimately be required to determine whether map-guided implantation translates into improved clinical outcomes and whether the map could eventually support decision-making for multiple plausible implantation options (including foregoing implantation altogether if all options lead to high non-responder likelihood).

### Limitations

Several limitations should be acknowledged. The small sample size retained after distance-based optimization (n=7) limits the generalizability of the 100% accuracy at this stage, but the score is nevertheless confirmed through multicenter validation. The limited sensitivity in the validation cohort (55%) likely reflects residual spatial mismatch between iEEG and RNS electrodes. This performance is expected to be substantially increased by a better proximity between the two types of electrodes, as would be the case with prospective map-guided implantation along iEEG trajectories. The imbalance between responders (n=31) and non-responders (n=18) may have influenced classification performance, though this concern is mitigated by use of balanced accuracy. Moreover, the reported imbalance in our three-center cohort is consistent with the expected rates of responders and non-responders reported in previous studies ^6^. The retrospective design precludes causal inference regarding the impact of lead placement on outcomes; whether map-guided implantation prospectively translates into improved clinical outcomes remains to be established in dedicated trials. Although electrode type heterogeneity across centers may have introduced variability in connectivity estimates, the generalization of the score across centers supports its robustness. Stimulation parameters (e.g., frequency, current amplitude) were not systematically controlled across patients, and may independently influence clinical outcomes. Accordingly, the spatial efficacy maps presented in Figure 3 should be interpreted as reflecting the combination of lead location and delivered stimulation, rather than location alone. Whether regions identified as suboptimal could yield better outcomes under different stimulation parameters remains an open question. Finally, the approach is not applicable to RNS candidates who did not undergo pre-implantation iEEG.

### Perspectives

Future prospective studies should assess whether map-guided RNS implantation translates into improved clinical outcomes, ultimately establishing the map as a standard component of pre-surgical planning. Additionally, combining the present connectivity-based approach with complementary biomarkers, such as post-implantation markers of network adaptation ^34^, may further refine patient selection and optimize stimulation strategies. The current framework could also be adapted to other biomarkers for an easy clinician-friendly visualization at the individual patient-level. More broadly, if replicated and prospectively validated, this framework could ultimately reduce the need for prolonged ictal monitoring: spatial efficacy maps derived from short interictal iEEG recordings could inform RNS lead placement, potentially streamlining the pre-surgical evaluation toward a more efficient, seizure-independent planning paradigm.

## Conclusion

In summary, iEEG functional connectivity predicts individual RNS response, and its performance is critically dependent on the spatial proximity between iEEG and RNS electrodes. Leveraging this spatial dependency, we derived patient-specific 3D maps of predicted RNS efficacy that could guide lead placement toward favorable network regions. These findings open a promising avenue toward network-informed, individualized RNS surgical planning.

## Data Availability

Code for the patient-level implementation of the biomarker will be made available following publication.

